# Adaptive forecasting of antiretroviral therapy demand using machine learning in India’s national HIV programme

**DOI:** 10.64898/2026.08.25.26361170

**Authors:** Muskaan Chugh, Bhavesh Neekhra, Manish Bamrotiya, Steven J. Clipman, Debayan Gupta

**Affiliations:** Department of Computer Science, Ashoka University, Sonipat, Haryana, India; Department of Medicine, Division of Infectious Diseases, Johns Hopkins University School of Medicine, Baltimore, Maryland, United States of America

## Abstract

Antiretroviral therapy (ART) stock-outs interrupt treatment, increase the risk of virologic failure and drug resistance, and erode the population-level benefits of viral suppression. India’s National AIDS Control Organization (NACO) manages one of the world’s largest public ART programmes, where regimen transitions, evolving formulations, changing treatment guidelines, and procurement-driven fluctuations in drug consumption complicate forecasting. We developed an end-to-end, regimen-specific forecasting workflow to support procurement planning during such periods of instability.

We analyzed monthly national ART consumption data from January 2013 through December 2024. A privacy-preserving synthetic dataset was used for pipeline development, followed by final evaluation on real national consumption time series. We compared three model classes, comprising six models: (1) classical models (Holt–Winters and ARIMA), (2) transformer models (TimesFM, which is a large pre-trained time-series foundation model, and its variant with logarithmically transformed values), and (3) hybrid models (two variants of a hybrid ARIMA-TimesFM residual model).

While the forecast horizon of 18 months remained constant, the train-test period varied across real and synthetic data, as real data were only available until February 2024. For synthetic data, models were trained through June 2023 (test window was July 2023-December 2024), while for real data, models were trained through August 2022 (our test window was September 2022-February 2024). We reported signed percentage deviation to preserve whether models tended to over-predict or under-predict, and selected models by the smallest absolute deviation. We then derived a regimen-specific model-error buffer, applied only to held-out under-prediction, and deployed the workflow through a no-code dashboard.

Forecasting performance was determined using signed percentage deviation (SPD), wherein positive deviation represents under-prediction and negative deviation represents over-prediction. Performance varied across regimens, indicating that no single approach was best-performing for all formulations. On synthetic benchmark data, the smallest absolute deviations ranged from 0.39% for adult AZT+3TC to 5.37% for adult DRV/r.

On real consumption data, classical methods remained competitive for some series, whereas transformer and hybrid models produced better predictive outcomes for others. For instance, for adult AZT+3TC, the Hybrid 70th percentile achieved an SPD of −2.02%, in contrast to the error range of [−15.7, 8.87] for other models. For adult RTV booster, Holt–Winters achieved an SPD of 1.68%, in contrast to the error range of [14.94%, 17.25%] for the other models.

Several formulations, particularly low-volume and transition regimens, nevertheless remained difficult to forecast accurately, underscoring persistent operational uncertainty. This was especially evident across the three pediatric regimens, where all models deviated systematically in the same direction — a more concerning pattern than mere error magnitude alone. For pediatric ABC+3TC, all models over-predicted within a narrow band of [−82.74, −67.43], while for pediatric AZT+3TC and LPV/r 100/25, all models under-predicted, with ranges of [24.93, 63.73] and [18.32, 52.07], respectively.

These findings support a portfolio approach to forecasting in national HIV programmes. Rather than replacing established public-health procurement systems, regimen-specific model selection, directional error reporting, and cautious model-error buffering can strengthen decision support during regimen transitions and other periods of unstable demand.

**Author summary:** Reliable demand forecasting for HIV medicines is essential to prevent stock-outs that interrupt treatment and compromise patient outcomes. In India’s public ART programme, procurement planning has traditionally drawn on past consumption. This approach performs reasonably well for mature, high-volume regimens, but becomes less reliable when national treatment guidelines change, new formulations are introduced or scaled up, or patients shift between regimens.

We used national ART consumption data from India to compare several forecasting approaches, including classical time-series models, a zero-shot foundation model, and a hybrid residual model. No single method performed best across all drug regimens; performance depended on the specific regimen being forecast. Classical models performed competitively on some relatively stable series, whereas others benefited from more flexible methods. We also developed a practical method for translating model output into procurement recommendations through a regimen-specific model-error buffer, and implemented the workflow in an accessible dashboard.

Our findings suggest that forecasting in national HIV programmes should be framed as a regimen-specific decision problem, rather than a search for one universally best model. This strategy could improve procurement planning during periods of programmatic change and help protect continuity of HIV treatment.

## Introduction

India continues to bear one of the world’s largest HIV burdens. National estimates for 2023 indicated approximately 2.6 million people living with HIV and about 66,400 new infections annually [1], in contrast to the estimates for 2021, which reported approximately 2.4 million people living with HIV and about 63,000 new infections annually [2]. Sustained progress towards the UNAIDS 95–95–95 targets requires reliable diagnosis, treatment access, and durable viral suppression [5]. Uninterrupted ART supply is essential for individual and population health – treatment interruptions are associated with subsequent treatment failure, and supply-chain failures contribute to HIV drug resistance in low- and middle-income settings [8, 9].

NACO administers a large, free public-sector ART programme, which expanded from 620 ART centers serving approximately 1.4 million people living with HIV in 2021 [4] to 725 ART centers providing lifelong treatment to approximately 1.6 million people as of June 2023 [3]. At this scale, forecasting and procurement are core programmatic functions, not merely logistics tasks. NACO’s own supply-chain guidance places uninterrupted commodity availability at the center of programme performance and uses explicit max–min inventory control, safety stock, and emergency order thresholds to prevent stock-outs and overstocking [20]. Forecasting accuracy therefore affects not only procurement efficiency, but also clinical outcomes, because forecast errors can propagate into service interruptions with direct patient-level consequences.

Forecasting ART demand is particularly difficult during regimen transition. WHO’s transition to Dolutegravir (DTG)-based regimens altered first-line treatment patterns across many national HIV programmes, and India similarly incorporated DTG into national guidance [4, 10]. Differentiated service delivery, including less frequent clinical visits and longer refill intervals for stable patients, can also alter dispensing patterns even when the number of patients on treatment remains unchanged [17, 18]. Pediatric formulations add further complexity because programmes must manage changing weight bands, product availability, and legacy stock already in the pipeline [11].

These features limit simple extrapolation from past consumption. Classical time-series models remain attractive because they are transparent and straightforward to implement, but they may struggle when structural breaks and nonlinear residual behavior are prominent. Hybrid models seek to combine interpretable statistical baselines with data-driven residual correction, and large pre-trained time-series foundation models such as TimesFM have been proposed as zero-shot forecasters capable of generalizing across previously unseen domains [12, 15]. Yet the comparative utility of these approaches for a national HIV supply chain, particularly under real programme conditions in a low- and middle-income country, remains incompletely characterized.

This study pursued three objectives. First, we characterized the current ART dispensing and procurement context, focusing on where demand forecasting can support existing public-health inventory workflows. Second, we evaluated forecasting approaches for drug procurement planning using national ART distribution series spanning adult and pediatric regimens with distinct demand profiles. Third, we translated these forecasts into actionable procurement guidance through a directional model-error buffer and a user-oriented, no-code dashboard for programme administrators and decision-makers. Rather than replacing existing inventory control mechanisms, our work aims to strengthen the forecasting inputs that inform them.

## Related work

Forecasting in healthcare and public-sector supply chains has traditionally relied on statistical models such as exponential smoothing and ARIMA, which remain common baselines in healthcare and medicine demand forecasting because they are relatively interpretable and can be applied to short historical series [13]. Forecasting ART demand poses additional challenges because demand reflects not only epidemiological burden but also treatment guideline changes, regimen transitions, stock availability, programmatic scale-up, and pediatric/adult regimen differences. These factors are especially relevant in HIV programmes, where treatment guidelines and preferred regimens have changed over time, with distinct considerations for adult and pediatric ART formulations [14].

Hybrid approaches have been used to improve performance when time series contain both linear and nonlinear structure, and healthcare-demand applications have reported gains from combining statistical baselines with adaptive residual correction [12]. More recently, foundation models for time-series forecasting have extended this work by using large-scale pre-training to support zero-shot prediction across domains; TimesFM is a prominent example [15].

Other relevant literature concerns inventory policy rather than forecasting alone. WHO’s procurement and supply management guidance emphasizes that forecasting is only one component of commodity security and should be interpreted alongside stock on hand, lead time, review period, safety stock, and early-warning indicators for stock-outs and overstocking [19]. NACO’s supply-chain SOP follows the same logic through an explicit max–min system, defined emergency order points, and routine stock review intervals [20]. This study sits at the interface of this literature: it evaluates alternative forecasting models, but frames them as decision-support inputs to a broader public-health supply chain.

## Methodology

### Study design and data sources

We developed an end-to-end forecasting workflow for national ART demand using monthly consumption data from India’s public HIV programme. The development process had two stages. First, we created a synthetic benchmark dataset to debug the modeling pipeline under privacy-preserving conditions and simulate known sources of volatility. Second, we applied the same workflow to real national aggregate ART consumption data received from NACO.

The analysis focused on eight regimen-specific time series spanning adult and pediatric formulations: ABC+3TC (adult and pediatric), AZT+3TC (adult and pediatric – serving as the backbone for ZLN/ZLE), ATV-based adult regimens, DRV-based adult regimens, Ritonavir adult use, and LPV/r 100/25 pediatric use. These series were chosen because they represent a mix of stable, lower-volume, pediatric, and transition-sensitive formulations relevant to national procurement.

**Table 1.**
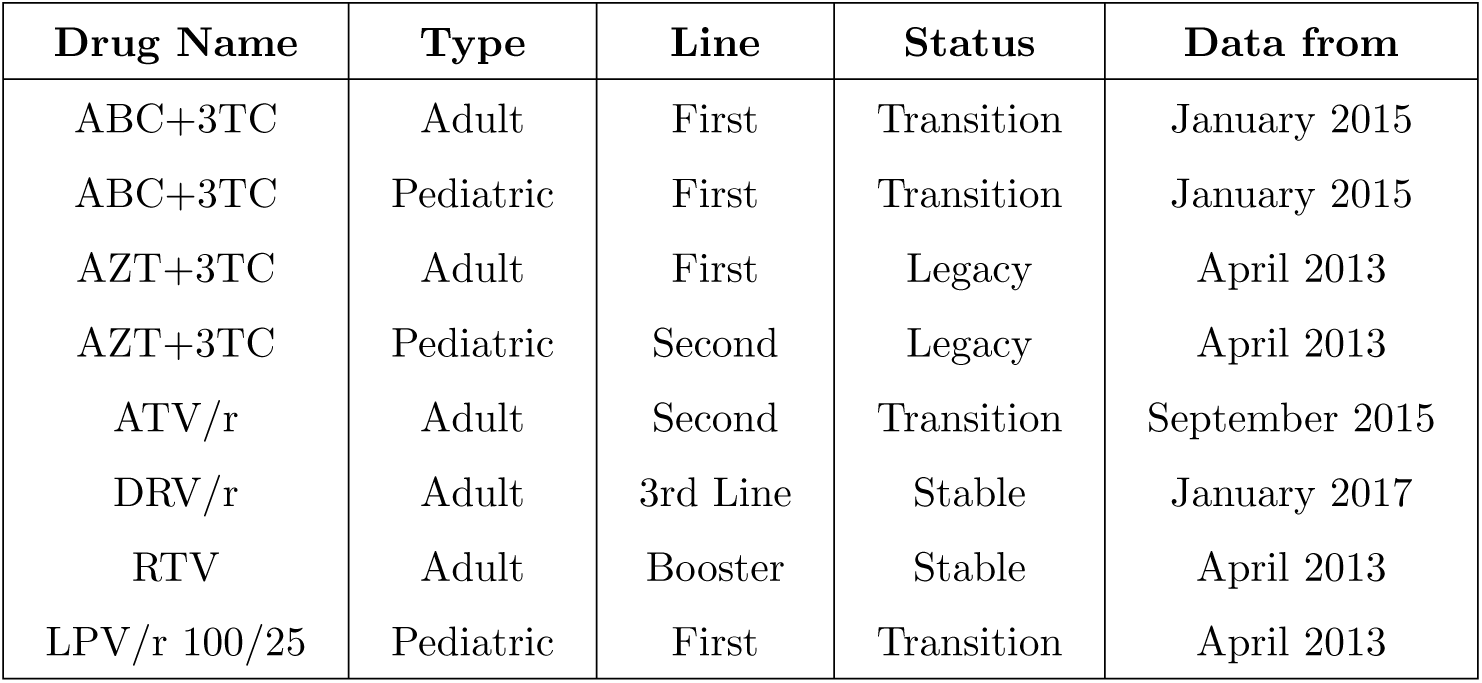
Details about the drugs under study.

| Drug Name | Type | Line | Status | Data from |
| --- | --- | --- | --- | --- |
| ABC+3TC | Adult | First | Transition | January 2015 |
| ABC+3TC | Pediatric | First | Transition | January 2015 |
| AZT+3TC | Adult | First | Legacy | April 2013 |
| AZT+3TC | Pediatric | Second | Legacy | April 2013 |
| ATV/r | Adult | Second | Transition | September 2015 |
| DRV/r | Adult | 3rd Line | Stable | January 2017 |
| RTV | Adult | Booster | Stable | April 2013 |
| LPV/r 100/25 | Pediatric | First | Transition | April 2013 |

Based on the National Guidelines for HIV Care and Treatment 2021 [4]

We limited our analysis to these eight regimens due to insufficient historical data for several newly introduced regimens. Dolutegravir-based combinations, including TLD (Tenofovir/Lamivudine/Dolutegravir), were not included as the transition to these regimens is ongoing and the available national consumption series are too short for the models evaluated here; their inclusion will become tractable as programme data accumulate.

### Synthetic data generation

Granular patient-level consumption data in public health are often scarce and subject to stringent confidentiality protocols, requiring a structured approach to data handling. We therefore developed and validated the pipeline on a synthetic dataset that reproduced the statistical properties and structural features of the real-world data before applying it to national-aggregate programmatic records, which reflect drug consumption volumes rather than individual patient demand. The synthetic dataset serves as a development and validation scaffold for the forecasting pipeline and does not constitute the evidentiary basis for the conclusions of this study.

#### Data source for synthetic baseline

To construct the synthetic dataset for our forecasting task on ART demand, we drew on publicly available programme statistics from India’s NACO and World Health Organization (WHO). Nine annual anchor points were established for fiscal years 2015-16 through 2023-24 using NACO-published line-wise ART totals – the number of patients on first, second, and third-line regimens – disaggregated by adults and children living with HIV (CLHIV) [21, 23–28]. Adult and pediatric patient counts were derived separately: adult counts were obtained directly from line-wise totals, whereas pediatric counts were estimated using the CLHIV-to-PLHIV ratio from India HIV Estimates as an anchor for the two observed fiscal years (2015-16, 2016-17) [30], with subsequent years estimated using a constant child-to-adult coverage ratio (k = 0.881) derived from those anchors.

Regimen-level shares were then applied to line-wise totals for each anchor year. Shares were not assumed arbitrarily but grounded in documented NACO guideline transitions: the 2013 guidelines established AZT+3TC as the preferred first-line regimen [31]; the 2017 guidelines displaced it in favor of TDF-based regimens, retaining AZT+3TC only for patients with documented TDF contraindications (estimated 5–6% in Indian cohorts, predominantly renal insufficiency) [33, 38]; and the 2021 guidelines introduced TLD (TDF+3TC+DTG) as the new preferred first-line, further marginalizing AZT+3TC [4].

ATV/r was the dominant second-line protease inhibitor (PI) through the early period, progressively displaced by DRV/r following its 2017 guideline introduction [33]; DRV/r maintained a share of 1.0 in the third-line treatment throughout, as it was the sole PI used in India’s public programme at that level [4]. Third-line patient counts were estimated by interpolating the third-to-second-line ratio from 0% (2016-17) to 4.63% (2019-20) [22, 23].

This yielded annual patient counts for eight series: ABC+3TC adult, AZT+3TC adult, ATV/r adult, DRV/r adult, ABC+3TC pediatric, AZT+3TC pediatric, LPV/r 100/25 pediatric, and RTV booster. RTV booster patient counts were not estimated independently; they were derived as the sum of patients on all three boosted regimens (ATV/r adult, DRV/r adult, LPV/r 100/25 pediatric), as ritonavir is dispensed as a fixed co-administration in each case [4, 33].

#### Interpolation and real-world simulation

Monthly patient counts were generated by linear interpolation between the nine fiscal-year anchor points (each anchored to the March value of the respective fiscal year), producing a continuous series from August 2015 to March 2024, covering 104 months.

Patient counts were then converted to monthly tablet consumption by multiplying by a regimen-specific dose-per-month figure derived from WHO weight-band dosing guidelines [32, 39]. Adult doses were: ABC+3TC 30 tablets/month (600/300 mg once daily), AZT+3TC 60 tablets/month (300/150 mg twice daily), ATV/r 30 tablets/month, DRV/r 60 tablets/month, and RTV booster 30 tablets/month (100 mg once daily). Pediatric doses were computed as weighted averages across WHO weight bands [39], using weight-band distributions informed by IeDEA global weight-for-age data [34] and India-specific age-at-diagnosis cohort data (median age *∼*10 years, 73% above 5 years) [35, 36].

One formulation-level change was modeled explicitly: Following WHO pre-qualification in April 2022, the 120/60 mg ABC+3TC dispersible tablet replaced the 60/30 mg formulation, halving the tablet count per dose and reducing pediatric ABC+3TC dose-per-month from 138 to 69 tablets [37].

Phase-specific Gaussian noise and discrete spike events were superimposed on each interpolated series to simulate real-world reporting variability. Noise was scaled to each regimen’s operational volatility: adult ABC+3TC noise declined from 2.5% (2015–2018) to 1.5% (2023–2024), reflecting programme maturation, while RTV booster was assigned a flat 4% noise rate across all phases given its dependence on co-administered PI regimens rather than direct prescription.

Spikes were sized and placed to mimic procurement surges, policy-driven reporting corrections, and supply disruptions; COVID-19-era disruptions (2020-2021) were represented through targeted negative deviations consistent with documented impacts on HIV service delivery [7, 8]. The synthetic benchmark was used exclusively for pipeline development and model comparison under controlled but clinically plausible conditions; it was not the primary evidentiary basis for the study’s conclusions. Generation parameters for all eight series are provided in the accompanying code repository.

#### Limitations of the synthetic benchmark

The synthetic dataset has several limitations that should be noted. First, monthly values between annual anchor points are products of linear interpolation; no monthly programme data are publicly available against which intra-year variation could be validated [21, 22, 24].

Second, the adult-to-pediatric patient split for fiscal years 2017-18 onward is estimated using a constant child-to-adult coverage ratio (k = 0.881) derived from only two observed anchor years (2015-16 and 2016-17) [21, 22]. This ratio was rising across those years, and holding it fixed produces a conservative lower bound on children on ART; the true split is unverifiable from public sources [30].

Third, AZT+3TC adult consumption is likely over-estimated: the size of the residual first-line AZT+3TC cohort following the 2017 TDF transition cannot be precisely determined from publicly available data, and the regimen shares used — while grounded in published TDF contraindication rates (estimated 5–6% in Indian cohorts) [38] — carry inherent uncertainty.

Fourth, a sharp discontinuity in India’s published CLHIV estimates between 2016–17 and 2017–18 (133,000 to 61,000) reflects a Spectrum model revision rather than a real demographic change, producing an artefact in the pediatric series around that period [30].

Fifth, pediatric weight-band distributions used in dose calibration are drawn from global IeDEA data [34]; India-specific distributions are not publicly available and may differ. These limitations are intrinsic to the use of a synthetic benchmark constructed from aggregate public data and do not affect the validity of the forecasting methodology evaluated against it.

### Real-world ART data and preprocessing

The primary analysis used real weekly NACO programme data spanning 2013–2024, depending on regimen availability. These data were reported by ART and Link ART facilities and arrived in multiple file formats, with naming conventions that varied across calendar periods. We harmonized regimen names across reporting eras, standardized state and union-territory identifiers, and aggregated weekly data to monthly national series. When multiple weekly observations were recorded within a calendar month, we used the most recent weekly observation as the monthly representative value. This approach reflects the structure of NACO’s reporting system, in which facilities record cumulative end-of-period consumption figures. The final weekly entry within a month therefore captures the most complete consumption record for that period. Months with no recorded observations were addressed through linear interpolation.

We addressed isolated missing monthly values through interpolation and harmonized reporting structures across three broad data eras (2013–2018, 2018–2022, and 2022–2024). The final modeling unit was the national monthly consumption volume per regimen, chosen to match the level at which central procurement decisions are most directly made.

### Forecasting models

We evaluated three core models and three additional variants or combination models derived from two of the core models. These can be broadly classified into three classes – classical time-series models, modern transformer models, and hybrid models. To facilitate reproducibility, we provide further implementation details in Appendix A.

#### Classical time-series models

1. **ARIMA:** This is a classical statistical model used to forecast time series data. It captures the linear dependencies in the data, including trends (auto-regressive), any differencing needed to make the series stationary (integrated), and short-term correlation of the forecast errors (moving average). It serves as a standard benchmark for stable, well-behaved time series. The integration order was determined via the Augmented Dickey–Fuller (ADF) test, which confirmed *d* = 1 across all eight series. Seasonal decomposition confirmed no meaningful seasonal component across any regimen (seasonal strength range: 0.035–0.264), and the model was specified accordingly.
2. **Holt–Winters:** This is a classical method particularly effective for time series data that exhibits both a clear trend (for example, continuously increasing demand) and seasonality (for example, predictable spikes every 12 months). It generates forecasts by using exponential smoothing for past observations and weights the most recent data more heavily. Additive trend and seasonality components were selected on the basis that seasonal amplitude did not scale consistently with the trend level across the evaluated series. Multiplicative components were not compared in the present study.

#### Modern transformer models

1. **TimesFM**: This is a large-scale, transformer-based model (developed by Google DeepMind) pre-trained on a massive, diverse corpus of time series. It operates in a zero-shot configuration, meaning it generates a forecast simply by being given historical data as context, without requiring any task-specific training or fine-tuning on the ART consumption data. It tests the transfer learning capability of deep learning models in the domain.
2. **TimesFM with Log-Transformation:** This approach uses the same TimesFM model after logarithmically transforming the original monthly demand data – a common preprocessing step used to stabilize the variance of the series and make the data distribution more nearly normal. We applied this transformation to potentially improve model performance on volatile series. The final forecasts are transformed back to the original scale.

#### Hybrid ARIMA-TimesFM models

This architecture is a two-stage hybrid approach designed to combine the strengths of both models. First, the ARIMA model provides a baseline forecast by capturing the linear, stable trends in the log-transformed data. Second, the actual errors (residuals) left by the ARIMA model are calculated. Crucially, these residuals are then passed to the zero-shot foundation model for forecasting. TimesFM is used here not just to predict a point error, but to model the uncertainty of the error, with the selected percentile defining the specific quantile of the error forecast.

In our analysis, we evaluated the 30th percentile, representing the lower bound of the forecast, and the 70th percentile, representing the upper bound. This allows the model to produce, not just a point forecast, but also a measure of its expected uncertainty, which is essential for determining safety stock.

#### Training setup

Each model used an 18-month forecast horizon, consistent with NACO’s supply-chain SOP, whereby stock sufficient for 18 months is procured during each procurement cycle [20]. The synthetic dataset spanned January 2015 through December 2024, with a testing window from July 2023 through December 2024. The available real data spanned January 2013 through February 2024, with a testing window from September 2022 through February 2024.

In the case of real data, the number of input observations varied by regimen, whereas the synthetic dataset contained 114 months of observations. Results from each model were stored for comparison to determine the ***best model*** for the chosen regimen. This design mirrored the practical decision problem faced by programme managers making forward procurement decisions with only historical data available.

### Evaluation framework

Forecasting models were evaluated on their ability to support procurement decisions within India’s national ART programme. Rather than seeking a single universally optimal model, the evaluation framework was designed to identify the best-performing approach for each regimen individually, reflecting the distinct demand dynamics of different formulations. Model accuracy was assessed using a directional metric that preserves the operational distinction between under- and over-prediction, and the selected model for each regimen was used to derive a procurement-oriented error buffer.

#### Accuracy metrics

To preserve the operational meaning of under- and over-forecasting, the main results tables report **signed percentage deviation** over the full holdout window:

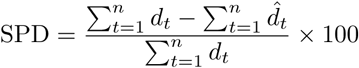

where *d_t_*is observed monthly consumption and *d^_t_* is forecast monthly consumption for month *t* in the holdout window. Positive values indicate under-forecasting (observed demand exceeded forecast), whereas negative values indicate over-forecasting (forecast exceeded observed demand).

Mean absolute percentage error (MAPE) and mean absolute error (MAE) were computed as supplementary descriptors of error magnitude:

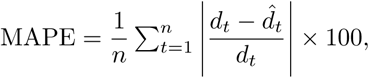

and mean absolute error (MAE) as

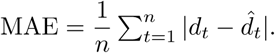

Neither metric was used in model selection; MAPE is known to be unreliable for low-volume or intermittent series [16] and is reported for descriptive context only.

#### Model selection criterion

Model selection for each regimen was based on the **absolute signed percentage deviation**,

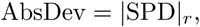

where SPD*_r_* is the signed percentage deviation for the selected model. This ensured that the model closest to zero was selected regardless of forecast direction.

NACO’s supply-chain SOP specifies three months of safety stock within a 6-month review cycle and a 3-month emergency order point [20]. A forecast error that consumes no more than half the safety stock buffer — reserving the remainder for supply-side uncertainty — implies a tolerable demand-side error of 1.5 months within a 6-month cycle, or approximately 25% of review-period demand. Applying a further conservative adjustment to avoid approaching the emergency order threshold yields an operational tolerance of roughly 15–20% for stable, high-volume regimens.

#### Model-error buffer

The model-error buffer for the forthcoming procurement cycle is derived from the model’s forecast deviation over the most recently completed procurement cycle, for which actual consumption data are available. This is a retrospective performance measure applied prospectively: the buffer captures how far the selected model deviated from observed demand in the prior period and applies that as a forward-looking safety margin. No future consumption values are used in its derivation.

The model-error buffer adjusts the total procurement forecast quantity and operates alongside — not in place of — NACO’s existing 3-month safety stock reserve, which is already embedded within the 18-month order window [18]. The two mechanisms address distinct uncertainty sources: safety stock covers supply-side variability (lead times, delivery delays), while the model-error buffer covers demand-side forecast error.

Forecasts were translated into procurement recommendations using a regimen-specific model-error buffer derived from the selected model’s under-prediction on the holdout data. Because only under-forecasting creates immediate stock-out risk, we defined the model-error buffer for regimen *r* as

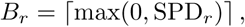

where SPD*_r_* is the signed percentage deviation for the selected model. If the selected model over-predicted overall (SPD*_r_* ≤ 0), no automatic buffer was applied, as over-prediction does not create immediate stock-out risk within NACO’s existing safety-stock framework [20]. The adjusted forecast was

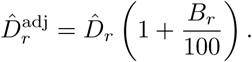

This buffer is intentionally simple and conservative. By basing the adjustment on historical under-prediction observed during validation, it provides an interpretable mechanism for incorporating model performance into procurement recommendations. The approach assumes that future model bias will broadly resemble that observed in the holdout period, and may therefore be less representative when under-prediction was driven by non-recurring events or when future demand dynamics differ substantially from the validation period.

As a supplementary forecasting input within NACO’s broader procurement framework — which already incorporates inventory targets, safety-stock requirements, periodic reviews, and operational oversight — these limitations do not undermine the buffer’s utility as a directional adjustment tool.

### Visualization and deployment tool

To support programme use, we implemented the forecasting workflow in a web-based, no-code dashboard. Users can upload updated data, select a forecast window, run regimen-specific forecasts, inspect the selected model for each regimen, and optionally apply the model-error buffer. To preserve usability, the default interface emphasizes forecasts and adjusted recommendations while allowing advanced users to inspect the underlying modeling steps.

## Results

### Performance on synthetic benchmark data

Performance varied across regimens, with no single model class dominating all series (Table 2). Based on the smallest absolute signed percentage deviation, Holt–Winters was selected for adult ABC+3TC (3.58%), adult ATV/r (0.52%), and adult DRV/r (5.37%). TimesFM was selected for adult RTV (0.86%) and pediatric AZT+3TC (0.48%), while log-transformed TimesFM was selected for adult AZT+3TC (0.39%). ARIMA was selected for pediatric ABC+3TC (1.93%) and pediatric LPV/r 100/25 mg (4.17%).

**Table 2.** Signed percentage deviation between forecast and observed consumption across drug regimens and forecasting approaches on synthetic ART benchmark data.

| Regimen | Type | Holt–Winters | ARIMA | TimesFM | TimesFM Log | Hybrid 30th | Hybrid 70th |
| --- | --- | --- | --- | --- | --- | --- | --- |
| ABC+3TC | Adult | **-3.58** | 12.42 | 12.79 | 11.60 | 15.64 | 10.17 |
| AZT+3TC | Adult | -6.18 | 13.73 | -1.44 | **-0.39** | 19.15 | 11.78 |
| ATV/r | Adult | **0.52** | -21.39 | 6.88 | -6.67 | -10.02 | -28.90 |
| DRV/r | Adult | **5.37** | -383.81 | 20.64 | -55.07 | -294.82 | -346.35 |
| RTV Booster | Adult | 11.21 | 2.00 | **0.86** | -22.15 | 14.19 | -2.09 |
| ABC+3TC | Pediatric | 74.48 | **1.93** | -74.95 | -52.47 | 11.37 | -3.23 |
| AZT+3TC | Pediatric | 28.63 | 13.08 | **-0.48** | -15.24 | 22.59 | 8.96 |
| LPV/r 100/25 | Pediatric | -5.98 | **-4.17** | -7.35 | -10.70 | 6.2 | -29.75 |

Table notes: Positive values indicate under-forecasting (observed consumption exceeded forecast); negative values indicate over-forecasting (forecast exceeded observed consumption). Boldface marks the selected model with the smallest absolute deviation for that regimen.

### Validation on real national consumption data

Performance on real NACO data was more heterogeneous and substantially more challenging than on the synthetic benchmark (Table 3). To contextualize model performance, we report the signed percentage deviation relative to programme projections over the forecast horizon. These projections have consistently over-estimated demand across all regimens with SPD ranging from −397.43% to −24%, a pattern consistent with conservative procurement planning intended to safeguard against stock-outs in a national supply chain.

**Table 3.** Signed percentage deviation between forecast and observed consumption across drug regimens and forecasting approaches on real national ART consumption data.

| Regimen | Type | Programme Projection | Holt–Winters | ARIMA | TimesFM | TimesFM Log | Hybrid 30th | Hybrid 70th |
| --- | --- | --- | --- | --- | --- | --- | --- | --- |
| ABC+3TC | Adult | -24 | **28.6** | 32.87 | 32.00 | 32.53 | 36.42 | 31.78 |
| ABC+3TC | Pediatric | -63.76 | -77.83 | -68.83 | -74.41 | -70.85 | **-67.43** | -82.74 |
| AZT+3TC | Adult | -34.40 | -3.22 | -4.88 | -15.7 | -13.2 | 8.87 | **-2.02** |
| AZT+3TC | Pediatric | -325.40 | **24.93** | 40.31 | 63.73 | 28.14 | 56.65 | 46.84 |
| DRV/r | Adult | -102.25 | **1.57** | -20.25 | 15.95 | 8.62 | -10.99 | -20.34 |
| ATV/r | Adult | -31.22 | -74.89 | -120.28 | -75.79 | **-61.83** | -103.82 | -118 |
| RTV | Adult | -98.74 | **1.68** | -14.21 | 17.25 | 6.53 | -5.2 | -14.94 |
| LPV/r 100/25 | Pediatric | -397.43 | 31.21 | 37.69 | **18.32** | 44.8 | 52.07 | 40.72 |

Table notes: Positive values indicate under-forecasting (observed consumption exceeded forecast); negative values indicate over-forecasting (forecast exceeded observed consumption). Boldface marks the selected model with the smallest absolute deviation for that regimen.

Among the evaluated models, classical methods performed competitively for select regimens: Holt–Winters achieved the smallest absolute deviation for adult ABC+3TC (28.6%), adult DRV/r (1.57%), adult RTV booster (1.68% with Holt–Winters), and pediatric AZT+3TC (24.93%). Other models outperformed the classical methods for the rest of the regimens, including pediatric ABC+3TC (67.43% with Hybrid 30th), adult AZT+3TC (2.02% with Hybrid 70th), adult ATV/r (61.83% with TimesFM Log), and pediatric LPV/r 100/25 (18.32% with TimesFM).

### Directional buffer recommendations and deployment

The directional interpretation of signed forecast deviation directly informed the buffer logic. For regimens whose selected model under-predicted over the holdout window, the dashboard applied a ceiling-based model-error buffer to the forecast. For regimens whose selected model over-predicted overall, no additional model-error buffer was automatically added. This prevented the system from inflating procurement quantities for series in which the chosen model already tended to overshoot demand.

Operationally, the dashboard displayed the selected model, signed deviation, and any resulting model-error buffer alongside the future forecast. Users could then either accept the model-derived recommendation or layer additional policy or managerial safety stock as needed.

## Discussion

The central finding of this study is that ART forecasting performance in India’s national programme was regimen-dependent: no single model class dominated the eight series evaluated, and the best-performing approach varied with the demand dynamics of each formulation. This result held on both synthetic and real data, although the magnitude of error differed markedly. It argues against adopting a single forecasting model for programme commodities and instead supports a portfolio approach in which model selection is tailored to each regimen.

The gap between synthetic and real-data performance warrants emphasis. On the synthetic benchmark, deviations for the best-performing model remained below 9% for all regimens. On real programme data, several formulations, particularly pediatric series and lower-volume regimens, showed deviations exceeding 30% even under the best available model. This divergence is expected. Synthetic series, however carefully parameterized, cannot reproduce the reporting irregularities, structural breaks, and policy-driven demand shifts that characterize live programme data. The practical implication is that synthetic benchmarks are useful for pipeline development, but should not be used to calibrate expectations for real-world forecasting accuracy.

The sources of this forecasting difficulty are identifiable. WHO’s transition to DTG-based first-line treatment and the introduction of new pediatric formulations require programmes to manage legacy stock while prescribing patterns shift [10, 11]. Differentiated service delivery can alter dispensing frequency without changing the number of patients on treatment [18]. These dynamics decouple historical consumption from future demand in ways that violate the stationary assumptions underlying classical extrapolation. The fact that several formulations remained difficult to forecast accurately, even with flexible models, underscores the limits of any purely statistical approach when the underlying demand-generating process is itself in transition.

These results also clarify the role of foundation models in public-health supply chains. TimesFM was evaluated in zero-shot mode – without task-specific fine-tuning on the ART dispensing data – because this represents the realistic deployment scenario for national programmes operating under data-use restrictions and limited computational infrastructure. Fine-tuning a foundation model requires both a sufficiently large labeled training set and ongoing retraining capacity, neither of which can be assumed in resource-constrained programme settings. Zero-shot evaluation therefore tests the more policy-relevant question: whether a pre-trained foundation model can contribute meaningful forecasting value without bespoke adaptation. In practice, TimesFM, applied in zero-shot mode, was not uniformly superior to classical methods [15]. Its performance varied by regimen and was sensitive to preprocessing decisions such as log transformation. Foundation models may therefore be most productively deployed within a regimen-specific selection framework rather than as default replacements for established methods. For stable, high-volume adult series, simpler models remained competitive and offered greater interpretability. For more volatile or low-volume formulations, flexible models – including the hybrid ARIMA-TimesFM residual approach – produced materially smaller deviations.

Beyond model comparison, this study makes a translational contribution by linking forecasts to procurement-oriented outputs through a directional model-error buffer and a no-code dashboard. The buffer applies only when the selected model under-predicted demand over the holdout window; over-forecasting, which does not create immediate stock-out risk, triggers no automatic upward adjustment. This directional logic is more operationally coherent than corrections based on unsigned error. The buffer is deliberately modest: it is a forecast-level overlay, not a substitute for NACO’s established max–min inventory policy, safety-stock norms, or emergency order points [20]. Forecasting should remain one input to a supply-chain decision process that also accounts for stock on hand, lead time, redistribution capacity, expiry risk, and procurement constraints [19].

Several limitations should be considered. The initial reliance on synthetic data, while necessary for privacy-preserving development, meant that the pipeline behavior could not be tested against real reporting artifacts until actual programme data were introduced. The national-level analysis aligns with central procurement but masks sub-national heterogeneity in both demand patterns and supply-chain capacity. Validation used a fixed 18-month holdout rather than a fully prospective programme evaluation, and the model-error buffer was deliberately simple, based on aggregate holdout under-prediction rather than a full stochastic inventory model. MAPE is known to be unreliable for intermittent or low-volume series [16]; we therefore emphasized signed deviation and interpreted percentage-based summaries with caution.

The most pressing next step is prospective evaluation: determining whether regimen-specific forecasting gains translate into fewer stock-outs, fewer emergency orders, and more stable continuity of care. Sub-national disaggregation, incorporation of leading indicators such as new-patient enrollment and regimen-switch rates, and integration with real-time inventory data are natural extensions. More broadly, these findings suggest that machine learning can strengthen public-health supply chains by improving the forecasting inputs on which those systems depend – precisely where consumption-based extrapolation is least reliable.

## Conclusion

Forecasting ART demand in a large national programme is not a single-model problem. National consumption data from India demonstrate that performance varies substantially by regimen and that regimen-specific model selection — rather than a uniform forecasting approach — is both feasible and operationally meaningful. Real programme data contained structural complexity not fully reproduced by the synthetic benchmark, and classical methods remained competitive for some mature adult series while flexible or hybrid methods delivered substantial improvements for lower-volume or transition-sensitive regimens.

These results support treating ART demand forecasting as a regimen-specific decision problem. Directional error reporting, under-prediction-based buffering, and deployment through an accessible dashboard provide a feasible translational pathway, but prospective evaluation is needed to determine whether improved forecasts translate into reduced stock-outs, fewer emergency orders, and greater continuity of treatment.

## Data Availability Statement

The real data underlying this study are national aggregate ART consumption records provided by India’s National AIDS Control Organization under a data-use agreement and are not publicly available. A validated synthetic dataset, the full analysis code, and dashboard source code are available on Zenodo (DOI: 10.5281/zenodo.21839042).

## Acknowledgments

We thank colleagues at the National AIDS Control Organization for technical discussions, for facilitating access to aggregate programme data, and for operational feedback during development of the forecasting workflow. We also thank the Johns Hopkins Gupta-Klinsky India Institute for the funding that made this work possible (GKII80064639).

## A Forecasting Implementation Details

### Classical time-series models

1. **ARIMA:** **Input:** Monthly consumption values transformed using log(1 + *x*) **Order search:** Non-seasonal orders *p, q* ∈ {0*,…,* 5}, with *d* = 1 confirmed by Augmented Dickey–Fuller test across all eight series **Seasonality:** None; seasonal decomposition confirmed no significant seasonal component across all eight series **Model selection:** Stepwise automatic ARIMA selection via AICc, initialized with *p* = 1 and *q* = 1 **Output:** Forecasts transformed back to the original consumption scale **Forecast horizon:** 18 months
2. **Holt–Winters:** **Input:** Untransformed monthly consumption values **Trend:** Additive **Seasonality:** Additive, with a seasonal period of 12 months **Initialization:** Estimated from the training data **Estimation:** L-BFGS-B optimization with brute-force initialization and bias correction **Output:** Non-negative forecasts on the original consumption scale **Forecast horizon:** 18 months

### Modern transformer models

1. **TimesFM**: **Input:** Untransformed monthly consumption values **Checkpoint:** google/timesfm-1.0-200m-pytorch **Inference:** Zero-shot forecasting without task-specific fine-tuning **Backend:** MPU (Apple Silicon), with a per-core batch size of 32 **Frequency:** Monthly time-series frequency category **Forecast horizon:** 18 months
2. **TimesFM with Log-Transformation:** **Transformation:** Monthly consumption values transformed using log(1 + *x*) **Checkpoint:** google/timesfm-1.0-200m-pytorch **Inference:** Zero-shot forecasting without task-specific fine-tuning **Backend:** MPU (Apple Silicon), with a per-core batch size of 32 **Back-transformation:** Forecasts transformed using exp(*x*) – 1 **Forecast horizon:** 18 months

### Hybrid ARIMA-TimesFM models

1. **Baseline model:** ARIMA fitted to log-transformed monthly consumption **Residual input:** The final 18 in-sample ARIMA errors on the logarithmic scale **Residual model:** Zero-shot TimesFM using the google/timesfm-1.0-200m-pytorch checkpoint **Quantiles:** The 30th- and 70th-percentile TimesFM residual forecasts were evaluated separately **Combination:** The ARIMA forecast and TimesFM residual forecast were combined on the logarithmic scale **Back-transformation:** Combined forecasts transformed using exp(*x*) – 1 **Forecast horizon:** 18 months

